# Monitoring SARS-CoV-2 circulation and diversity through community wastewater sequencing

**DOI:** 10.1101/2020.09.21.20198838

**Authors:** Ray Izquierdo-Lara, Goffe Elsinga, Leo Heijnen, Bas B. Oude Munnink, Claudia M. E. Schapendonk, David Nieuwenhuijse, Matthijs Kon, Lu Lu, Frank M. Aarestrup, Samantha Lycett, Gertjan Medema, Marion P.G. Koopmans, Miranda de Graaf

**Affiliations:** Department of Viroscience, Erasmus University Medical Center, Rotterdam, The Netherlands; KWR Water Research Institute, 3433 PE, Nieuwegein, The Netherlands; Usher Institute, University of Edinburgh, Edinburgh, UK; National Food Institute, Technical University of Denmark, Kongens Lyngby, Denmark; The Roslin Institute, University of Edinburgh, Edinburgh, UK

**Author notes:** These authors jointly supervised this work: Gertjan Medema, Marion Koopmans, Miranda de Graaf.

**Keywords:** sewage, wastewater-based epidemiology, SARS-CoV-2, Nanopore, Illumina

## Abstract

The current SARS-CoV-2 pandemic has rapidly become a major global health problem for which public health surveillance is crucial to monitor virus spread. Given the presence of viral RNA in feces in around 40% of infected persons, wastewater-based epidemiology has been proposed as an addition to disease-based surveillance to assess the spread of the virus at the community level. Here we have explored the possibility of using next-generation sequencing (NGS) of sewage samples to evaluate the diversity of SARS-CoV-2 at the community level from routine wastewater testing, and compared these results with the virus diversity in patients from the Netherlands and Belgium. Phylogenetic analysis revealed the presence of viruses belonging to the most prevalent clades (19A, 20A and 20B) in both countries. Clades 19B and 20C were not identified, while they were present in clinical samples during the same period. Low frequency variant (LFV) analysis showed that some known LFVs can be associated with particular clusters within a clade, different to those of their consensus sequences, suggesting the presence of at least 2 clades within a single sewage sample. Additionally, combining genome consensus and LFV analyses we found a total of 57 unique mutations in the SARS-CoV-2 genome which have not been described before. In conclusion, this work illustrates how NGS analysis of wastewater can be used to approximate the diversity of SARS-CoV-2 viruses circulating in a community.

## INTRODUCTION

Since the discovery of the Severe Acute Respiratory Syndrome 2 (SARS-CoV-2), the agent causing the coronavirus disease 2019 (COVID-19), in late December 2019^1^, the virus has caused more than 30 million confirmed cases and more than 950,000 deaths worldwide (September 18^th^ 2020, https://coronavirus.jhu.edu/). The virus belongs to the family *Coronaviridae*, genus *Betacoronavirus* and subgenus *Sarbecovirus*^2^. The impact of the virus and the interest to understand its origin and epidemiology have resulted in the sequencing of more than 105,000 genomes as of September 18^th^, 2020 (https://www.gisaid.org/)^3^. This has allowed the identification of groups of viruses based on their genetic diversity that can be related to geographical and temporal patterns^4^. Recently, based on the lineages assigned by Rambaut et al. (2020)^4^, Nextstrain has proposed a classification for SARS-CoV-2 strains based on the high prevalence (>20%) of signature mutations, persistence overtime and geographic spread (https://nextstrain.org/)^5^. Currently, Nextstrain classification divides SARS-CoV-2 diversity into five major global clades: 19A, 19B, 20A, 20B and 20C.

Although SARS-CoV-2 infects primarily respiratory tract tissues, it has been shown that the virus is able to efficiently replicate in the gastrointestinal tract, as evidenced by *in vitro* infection of enteroids^6^, presence of viral proteins in gastrointestinal epithelium biopsies^7^, and detection of infectious virus in stool samples^8^. The virus attaches to host cells by binding of the Spike (S) glycoprotein to its human receptor, the angiotensin-converting enzyme 2 (ACE2)^9,10^. ACE2 is a cellular membrane glycoprotein highly expressed in several organs such as the lungs, kidneys, heart, as well as the gut and infection has been confirmed in specific cells in or derived from each of these organs, including enterocytes^6,11^. Viral RNA is shed in feces of around 40% of the infected patients, often for longer periods than for nasal swabs, SARS-CoV-2 RNA detection in urine has been observed less occasionally (< 5% of infected patients)^12,13^. Nevertheless, the role of the fecal-oral transmission route in the epidemiology of the virus is still unknown^12^.

Due to the rapid spread of SARS-CoV-2, individual screening of clinical cases is challenging. This is even more difficult when studying the viral diversity on a population level, since a large number of clinical and non-clinical samples would need to be sequenced. Various reports have shown that it is possible to detect enteric viruses in wastewater, and also respiratory viruses such as Influenza A, SARS-CoV and SARS-CoV-2^14–19^. This has led to the recognition of the potential of wastewater-based epidemiology as a valuable tool to assess the spread of the disease at a community level. Recently, the Water Research Institute (KWR) in the Netherlands, as well as others, have demonstrated temporal correlations between SARS-CoV-2 RNA titers in sewage and the number of cases in a particular city or county, where the titers in sewage seem to correlate with the number of reported cases in the population, suggesting a potential role for sewage surveillance as an early warning tool^18,20–22^. Therefore, sewage testing is currently considered globally as an adjunct to patient-based surveillance, and has promise as an early warning indicator of increasing virus circulation. Enhanced surveillance is a key pillar of the current containment strategy aiming to control the spread of SARS-CoV-2 and includes frequent testing of people with mild symptoms, investigation of clusters of infection to identify possible common exposures, and monitoring of hospital and ICU admissions. Whole genome sequencing of SARS-CoV-2 directly from clinical samples has been developed as an additional tool, to provide information on diversity of circulating strains as a basis for cluster identification. Particularly in areas with minimal circulation, sequencing of viruses from patients can help to identify a possible source, provided that sufficient background sequencing has been done. So far, little work is done trying to correlate the SARS-CoV-2 diversity in sewage and patients^23,24^. Here we aimed to evaluate the potential of next generation sequencing (NGS) of SARS-CoV-2, from RT-PCR positive wastewater samples, to assess if they reflect the diversity of SARS-CoV-2 circulating within the population of the Netherlands and Belgium.

## RESULTS

### Correlation between RT-qPCR and the percentage of genome recovered by NGS

Previously, sewage samples were collected from different locations in The Netherlands and Belgium to investigate the levels of SARS-CoV-2 in sewage using RT-qPCR^18^. To further investigate the genetic diversity of SARS-CoV-2 a total of 55 wastewater samples obtained from 13 different locations in the Netherlands (48 samples) and 7 different locations in Belgium (7 samples) with Ct values of <36 were selected for whole genome sequencing using Nanopore sequencing. The samples covered a time span of 70 days (from March 25^th^ to June 3^rd^ 2020). Two samples (Franeker-92719 and AmsterdamWest-92852) were sequenced by Nanopore twice, while 24 samples were also sequenced by Illumina (Table 1). Four primers/probe sets targeting the N (N1-N3)^25^ and the E genes^26^ were used to evaluate the presence and concentration of SARS-CoV-2 in sewage samples as described previously^18^. All samples and their Ct values are shown in Table 1. The percentage of the genome covered by the assembly of Nanopore reads (>10X coverage per site) ranged from 0 to 99.2%. We observed an inverse sigmoidal correlation between the percentage of the genome assembled from Nanopore sequencing reads and the Ct values of both the N2 and the E primers/probe sets (Fig. 1). The inflection point (Ct value at which half of the genome can be obtained) for N1, N2, N3 and E primers/probe sets were Ct values of 34.6, 33.8, 33.2 and 32.5, respectively. No correlation was observed between Ct values and the percentage of the genome assembled from Illumina sequencing data (Supplementary Fig. S1).

**Table 1.**
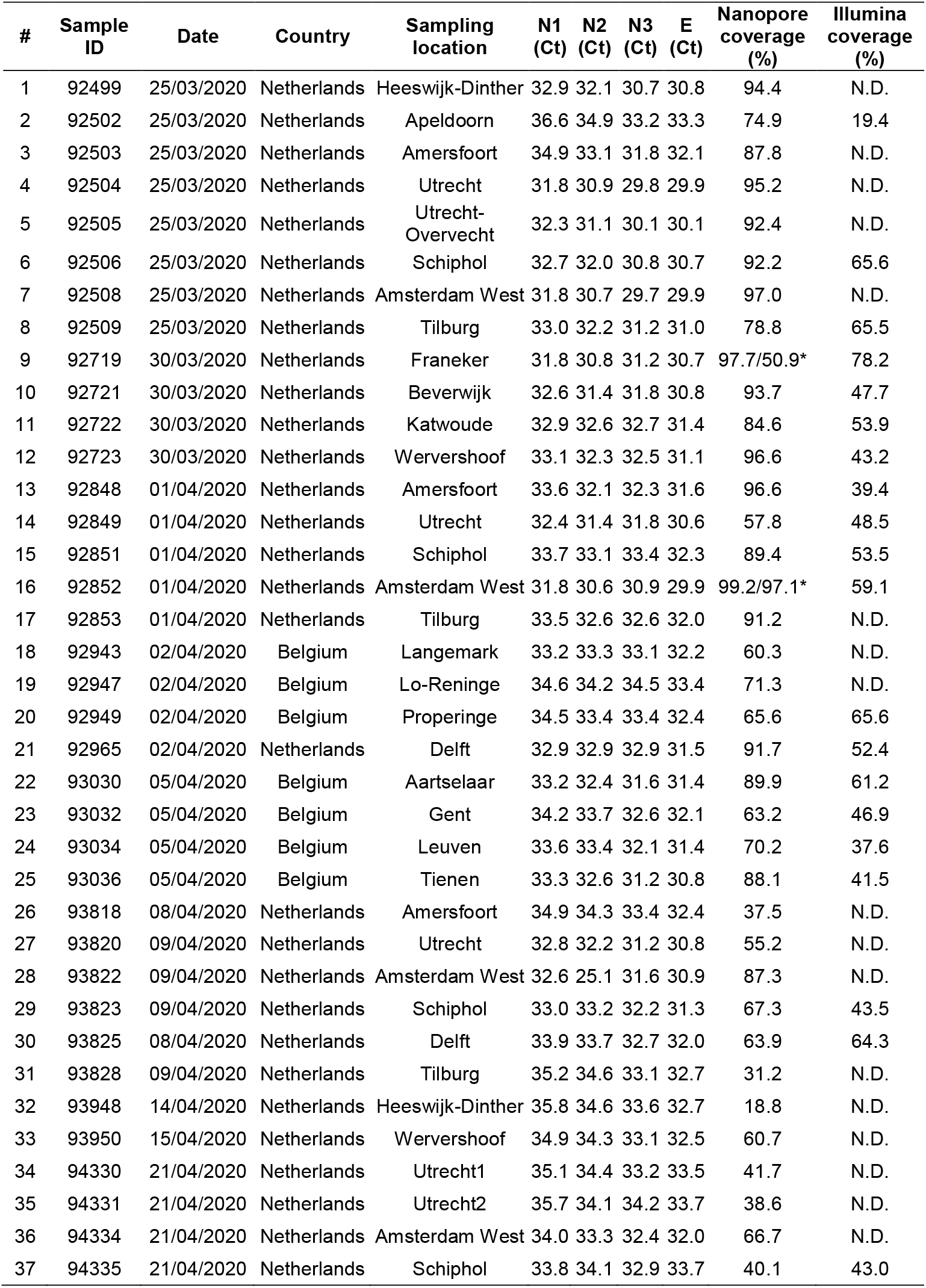
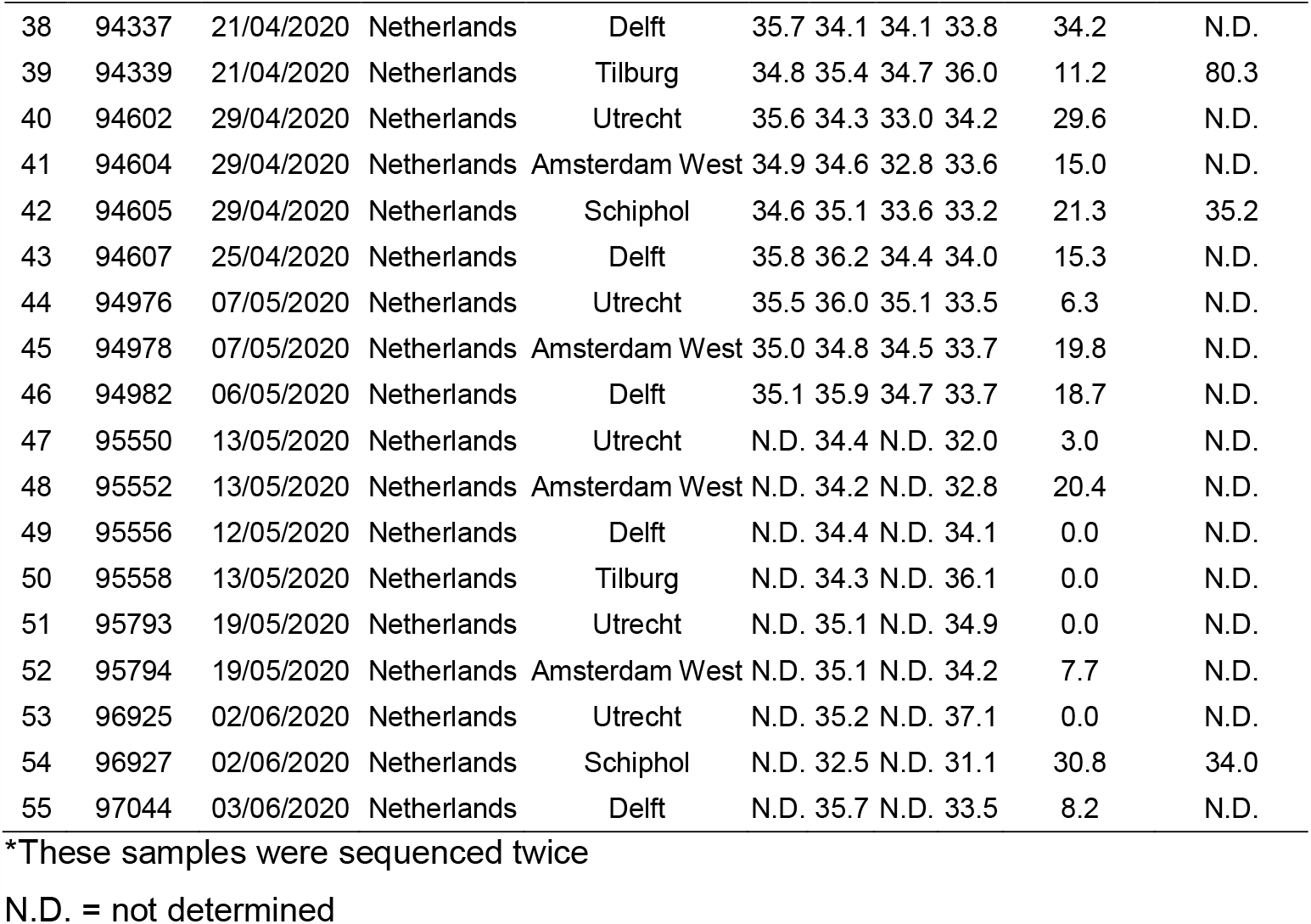
Overview of the SARS-CoV-2 wastewater samples sequenced during this study

**Fig 1.**
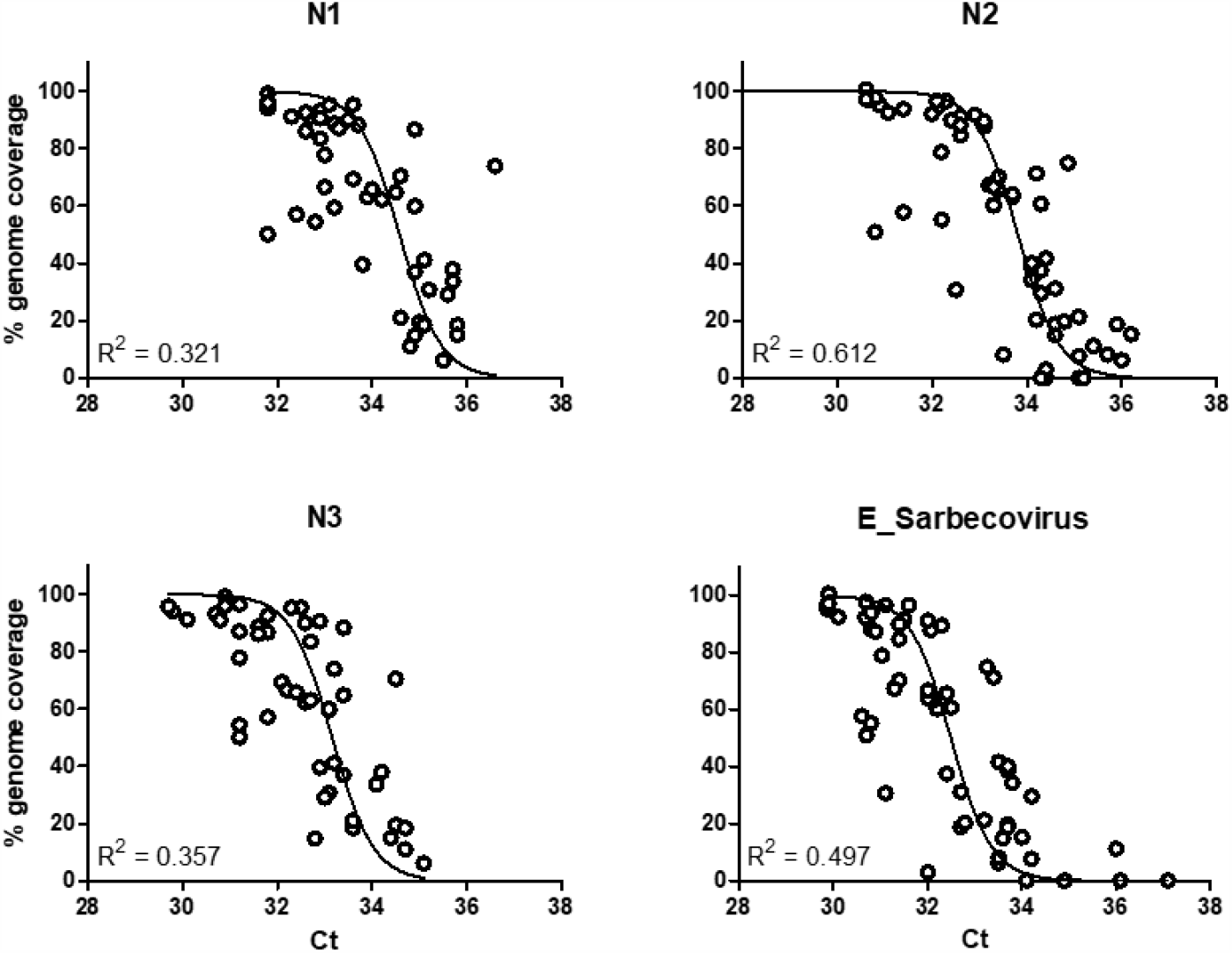
RT-qPCR cycle threshold (Ct) of SARS-CoV-2 RNA in sewage as determined by the N gene (N1-N3) and the E gene RT-qPCR assays against the percentage of the genome covered (> 10X) by Nanopore reads.

### Consensus sequences from sewage may reflect the most prevalent viruses in human populations

A total of 22 genomes with a coverage >75% of the genome were obtained from 20 samples, 20 from the Nanopore runs and 2 from the Illumina run. These consensus sequences were used to infer two maximum-likelihood (ML) trees, the first using all Dutch and Belgian sequences available in GISAID database and the second using a representative subset of the Global diversity of SARS-CoV-2 in GISAID database. In general, Dutch and Belgian sequences are grouped into the 5 clades (Fig 2a), where most of the sequences belong to clade 20A with 52.0% and 47.7% prevalence in Dutch and Belgian sequences, respectively. 19B and 20C are the less prevalent clades with 8.9% and 1.2% for Dutch sequences and 10.4% and 0.3% for Belgian sequences, respectively. Both trees showed that sewage samples were grouped within clades 19A, 20A and 20B (Figs 2a and b). None of the sewage consensus sequences were found within clades 19B or 20C. Some sewage samples clustered with sequences isolated from patients of the same region, such as sequences from sample Franeker-92719 (both, Illumina and Nanopore consensus sequences) and hCoV-19/env/Netherlands/HeeswijkDinther-92499-N/2020 (Fig 2a). There were 2 samples with 2 consensus sequences included in the phylogenetic trees (AmsterdamWest-92852 and Franeker-92719), which interestingly had a 2-mutation difference between consensuses sequences of the same sample in both cases (Supplementary Table S1). Despite this, consensus sequences from the same sample cluster within the same clade (Supplementary Figs. S2 and S3). Some sequences were found to be close to the root of the clades, probably due to the presence of multiple virus strains within one sample resulting in a combination of mutations in their consensus sequences (Supplementary Figs. S2 and S3).

**Fig 2.**
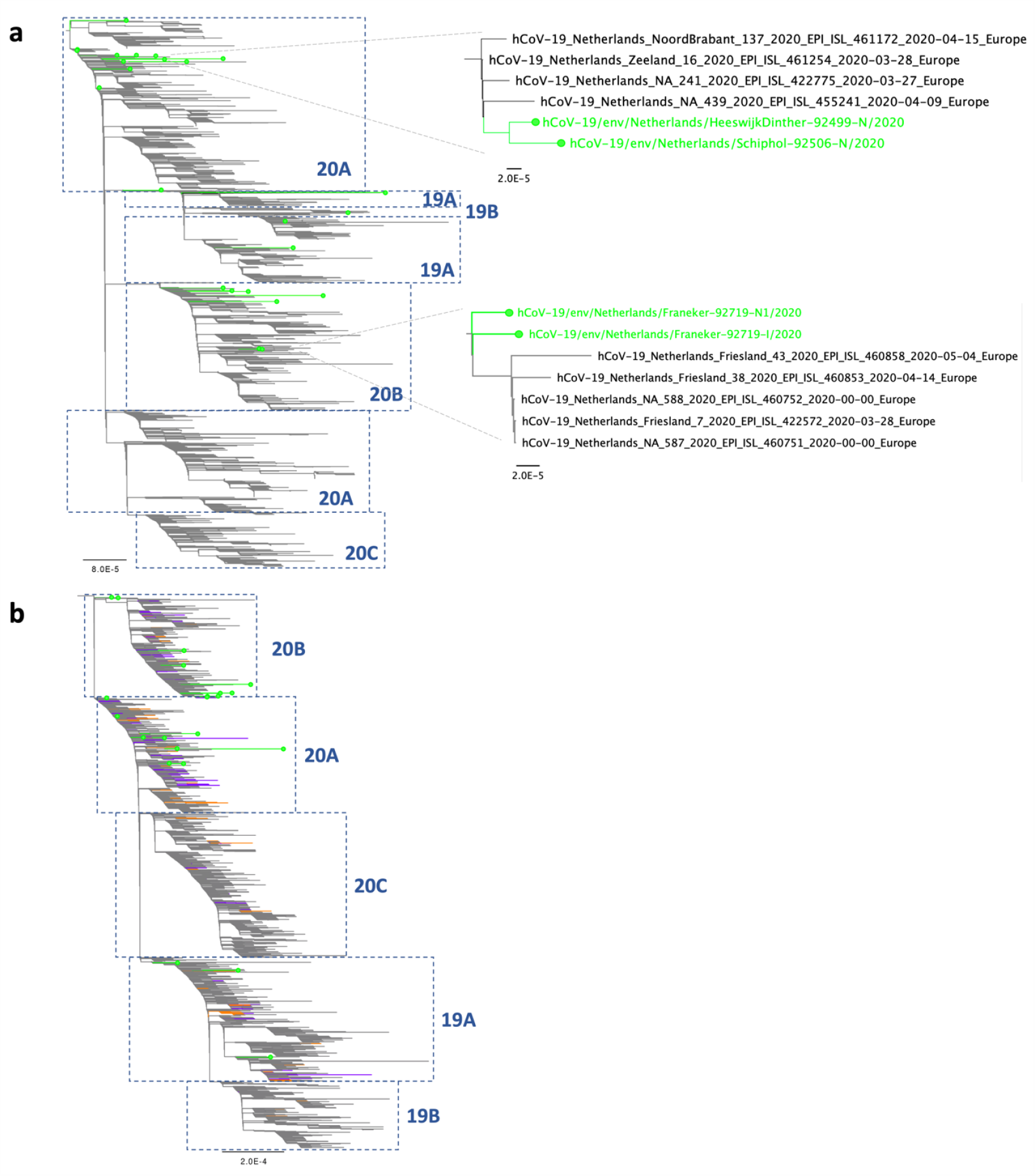
Phylogenetic analysis of SARS-CoV-2 genome consensus sequences detected in sewage samples in the Netherlands and Belgium, using a Dutch (a) and a Global subsample (b) dataset. Lines with dots in green indicate samples sequenced in this study. Clades (19A, 19B, 20A, 20B and 20C) were assigned using the Nextclade tool (https://clades.nextstrain.org/). For the Global subsample tree, samples in orange and purple indicate Dutch and Belgian sequences, respectively.

In order to associate specific mutations to particular a clade or cluster, all consensus sequences, including partial sequences, were compared to the Wuhan-Hu-1 reference sequence. A total of 145 single nucleotide polymorphisms (SNPs) compared to the Wuhan-Hu-1 reference sequence were detected in our dataset (supplementary Table S1). From these, 24 SNPs were present in more than one sequence. The maximum number of mutations in individual samples compared to the Wuhan-Hu-1 reference genome were 11 for hCoV-19/env/Netherlands/Amersfoort-92503-N/2020, hCoV-19/env/Netherlands/Delft-92965-N/2020 and hCoV-19/env/Netherlands/Schiphol-94335-N/2020 (supplementary Table S1). The presence of clade-defining mutations in the consensus sequence suggests the dominance of a certain clade within a sample, but assessing their presence can also be used to check for virus mixtures in a sample. Nextstrain has defined each clade by the presence of at least two linked mutations (https://nextstrain.org/). 19A is the root clade and contains the Wuhan-Hu-1 reference sequence. Both 19B and 20A emerged from 19A, where two and three linked mutations define these major clades, respectively: T28144C and C8782T for 19B; and C3037T, C14408T and A23403G for 20A. Clades 20B and 20C emerged from 20A, where the trinucleotide substitution GGG > AAC at positions 28881-28883 defines 20B, and the linked mutations C1059T and G25563T define 20C. Nucleotide substitution A23403G, a signature mutation of clades 20A, 20B and 20C, and that generates the D614G amino acid substitution in the S glycoprotein, was present in 83.6% (51/61) of the samples that were sequenced at this region (supplementary Table S1). The 20B signature mutation, GGG > AAC at positions 28881-28883, that results in the change of 2 amino acids, RG to KR, in the Nucleocapsid (N) at positions 203-204^27^ was present in 41.9% (18/43) of the sequences (supplementary Table S1). One of the two mutations defining clades 20C and 19B (C1059T and T28144C) were found in 2 and 3 consensus sequences, respectively. The regions containing the other clade-defining mutations (C25563T for 20C; and C8782T for 19B) were not sequenced at high enough coverage to determine a consensus sequence in these samples. The hCoV-19/env/Netherlands/Amersfoort-92503-N/2020 sequence contained a mix of the clade-defining mutations C1059T, T28144C and GGG28881-28883AAC, that define clades 20C, 19B and 20B, respectively. This indicates that the obtained consensus sequence is a combination of several different viruses and does not represent a single strain.

In addition to the clade-defining mutations, we detected 49 and 63 SNPs in sewage samples that were not present in either the Dutch (1544 sequences) or Belgian (888 sequences) GISAID datasets, respectively, but were present in the Global dataset (55074 sequences, as 8^th^ of July 2020), although with < 1% prevalence (supplementary Table S2). Moreover, we detected a total of 51 novel mutations present in sewage consensus sequences that were not previously reported (supplementary Table S2), of which 48 were supported by coverage above the set thresholds to be considered as high quality (coverage >30x for Nanopore; and coverage >5X and Phred score >30 for Illumina). Additionally, it is noteworthy to mention that some samples presented discrepancies between the consensus sequences obtained by Nanopore and/or Illumina sequencing. For example, sample AmsterdamWest-92852 was sequenced 3 times (twice with Nanopore and once with Illumina), in which 4 positions with discrepancies were found between the consensus sequences (Supplementary Table S1).

These differences were not due to an assembly error, since the alignment of the reads were manually checked and corrected for each discrepancy in every sequencing run. These differences were explained by the presence of two different nucleotides in the reads covering a particular position with varying percentages between sequencing runs, resulting in consensus sequences that could differ between each sequencing run. These results were likely caused by both or either the presence of multiple strains and low viral RNA titers in the samples, leading to an amplification bias during library preparation.

### Low frequency variant (LFV) analysis revealed both known and novel mutations for SARS-CoV-2

Given that sewage samples are likely to contain a mixture of SARS-CoV-2 strains, we decided to perform a variant analysis with Illumina data to determine whether LFVs were confidently present in a sample. Using a coverage > 50X, Phred score > 30 and a frequency threshold of > 10% as settings, we found a total of 21 positions with at least one sample containing major and minor variants (Table 2). From these, 14 mutations resulted in changes at the amino acid level (12 non-synonymous mutations and 2 deletions). Interestingly, 8 of these variants (4497C, 10514C, 11484T, 13046A, 16538_16540delATA, 16777T, 16823T and 28736A) are novel mutations that were not present in either the Dutch-Belgian (2432 sequences) or the Global (55074 sequences) GISAID datasets. The other 7 variants were present with low prevalence in both datasets (from 0.002% to 0.130%). The most prominent of these was the 28139A mutation, that was present in only 4 sequences worldwide, showing both a temporal (all detected in March 2020) and a regional signal (2 sequences from the Netherlands [EPI_ISL_422640 and EPI_ISL_422880], 1 from Denmark [EPI_ISL_444879] and 1 from Belgium [EPI_ISL_458209]). Finally, 4 variants (1440A, 11083T, 11109T and 24862G) were present at higher levels in both datasets (> 0.5%), where 11109T and 24862G are 28.5 and 14.3 times more prevalent in the Dutch dataset than in the Global dataset, and 45.0 and 35.5 times more compared with the Belgian dataset, respectively (Table 2).The other variants were present at similar frequencies in the Dutch, Belgian and Global datasets (Table 2).

**Table 2.** Summary of the Low frequency variants (LFV) detected in wastewater samples determined by Illumina sequencing.

| Position<sup>(a)</sup> | Sample | Mayor Variant | LFV | LFV freq (%) | Total Depth | Feature | Effect<sup>(b)</sup> | AA MV | AA LFV | NL freq (%)<sup>(c)</sup> | BE freq (%)<sup>(c)</sup> | Global freq (%)<sup>(c)</sup> |
| --- | --- | --- | --- | --- | --- | --- | --- | --- | --- | --- | --- | --- |
| 1440 | Netherlands/Schiphol-92506-I | G | A | 13.2 | 53 | ORF1a | NS | G | N | 1.619 | 4.167 | 1.903 |
| 3549 | Netherlands/Franeker-92719-I | GACCACTTA | - | 46.8 | 201 | ORF1a | Del | GPLK | E | 0.000 | 0.000 | 0.002 |
| 4497 | Netherlands/Beverwijk-92721-I | T | C | 42.6 | 479 | ORF1a | NS | I | T | 0.000 | 0.000 | 0.000 |
| 10514 | Netherlands/AmsterdamWest-92852-I | T | C | 12.5 | 1656 | ORF1a | NS | Y | H | 0.000 | 0.000 | 0.000 |
| 10933 | Belgium/Aartselaar-93030-I | C | T | 18.0 | 50 | ORF1a | SYN | P | P | 100.000 | 100.000 | 99.996 |
|  | Netherlands/Tilburg-94339-I | T | C | 11.1 | 63 | ORF1a | SYN | P | P | 0.000 | 0.000 | 0.004 |
| 11083 | Belgium/Properinge-92949-I | G | T | 12.1 | 58 | ORF1a | NS | L | F | 5.635 | 7.320 | 11.007 |
|  | Belgium/Aartselaar-93030-I | T | G | 26.4 | 129 | ORF1a | NS | F | L | 94.430 | 92.680 | 88.069 |
|  | Netherlands/Tilburg-94339-I | G | T | 12.7 | 150 | ORF1a | NS | L | F | 5.635 | 7.320 | 11.007 |
| 11109 | Netherlands/AmsterdamWest-92852-I | C | T | 48.3 | 230 | ORF1a | NS | A | V | 15.220 | 0.338 | 0.534 |
|  | Netherlands/Tilburg-94339-I | C | T | 21.2 | 66 | ORF1a | NS | A | V | 15.220 | 0.338 | 0.534 |
| 11484 | Netherlands/Beverwijk-92721-I | C | T | 44.0 | 84 | ORF1a | NS | A | V | 0.000 | 0.000 | 0.000 |
| 11494 | Netherlands/Franeker-92719-I | C | T | 13.5 | 104 | ORF1a | SYN | N | N | 0.000 | 0.000 | 0.002 |
|  | Belgium/Aartselaar-93030-I | C | T | 43.5 | 370 | ORF1a | SYN | N | N | 0.000 | 0.000 | 0.002 |
|  | Netherlands/Tilburg-94339-I | C | T | 13.8 | 247 | ORF1a | SYN | N | N | 0.000 | 0.000 | 0.002 |
| 13046 | Belgium/Aartselaar-93030-I | C | A | 36.7 | 98 | ORF1a | NS | P | T | 0.000 | 0.000 | 0.000 |
| 13426 | Belgium/Gent-93032-I | C | T | 22.6 | 115 | ORF1a | SYN | R | R | 0.000 | 0.000 | 0.038 |
| 16538 | Belgium/Gent-93032-I | - | ATA | 27.6 | 348 | ORF1b | Del | - | N | 100.000 | 100.000 | 100.000 |
| 16777 | Netherlands/Schiphol-92851-I | G | T | 30.2 | 404 | ORF1b | NS | V | F | 0.000 | 0.000 | 0.000 |
| 16806 | Netherlands/Tilburg-94339-I | C | A | 22.1 | 77 | ORF1b | NS | N | K | 0.000 | 0.000 | 0.016 |
| 16823 | Belgium/Aartselaar-93030-I | G | T | 12.0 | 192 | ORF1b | NS | G | V | 0.000 | 0.000 | 0.000 |
| 24862 | Netherlands/Katwoude-92722-I | A | G | 34.0 | 53 | S | SYN | T | T | 8.614 | 0.338 | 0.463 |
| 28115 | Netherlands/Delft-92965-I | T | C | 47.8 | 67 | ORF8 | SYN | I | I | 100.000 | 100.000 | 99.993 |
| 28139 | Netherlands/Tilburg-94339-I | C | A | 36.0 | 136 | ORF8 | SYN | S | S | 0.130 | 0.113 | 0.007 |
| 28375 | Netherlands/Tilburg-94339-I | G | A | 30.8 | 146 | N | SYN | G | G | 0.000 | 0.000 | 0.002 |
| 28394 | Netherlands/AmsterdamWest-92852-I | C | T | 31.5 | 54 | N | NS | R | W | 0.000 | 0.000 | 0.004 |
|  | Belgium/Properinge-92949-I | C | T | 16.7 | 60 | N | NS | R | W | 0.000 | 0.000 | 0.004 |
|  | Netherlands/Tilburg-94339-I | C | T | 13.6 | 191 | N | NS | R | W | 0.000 | 0.000 | 0.004 |
| 28736 | Belgium/Leuven-93034-I | A | G | 22.0 | 363 | N | NS | A | T | 100.000 | 100.000 | 100.000 |

(a) Positions are given with respect to Wuhan-Hu-1 (MN908947).
(b) NS = non-synonymous; SYN = synonymous mutation; Del = deletion
(c) Frequency of the LFV of the sample against GISAID database as up to July 8<sup>th</sup> 2020.

### Presence of specific LFVs may be associated to the presence of viruses belonging to a particular cluster

In addition to consensus sequence, LFV analysis is of importance to be able to identify potential local outbreaks from sewage. To try to associate the presence of a minor variant to sequences belonging to unique clusters, we mapped the 4 most highly prevalent LFVs onto both Dutch-Belgian and global subsample phylogenetic trees (Fig 3). This analysis indicated that for 3 variants (1440A, 11109T and 24862G) there were clear associations between the presence of the mutation and their clustering on the phylogenies (Fig 3). However, when one of these 3 variants was present as an LFV in a sewage sample the consensus sequence (blue arrows in Fig. 3) of this sample did not group with the cluster of clinical samples that contains the variant (magenta lines in Fig. 3). For example, 24862G variant in sample Tilburg-94339 was present in two unique clusters within clade 20A, while its consensus sequence (hCoV-19/env/Netherlands/Tilburg-94339-I/2020) was clustered within clade 20B (Fig 3 and Supplementary Figs. S2 and S3), suggesting the presence of both clades in this sample. Although mutation 11083T was most prevalent in clade 19A, it was also present scattered along the trees, indicating poor association with a particular clade.

**Fig 3.**
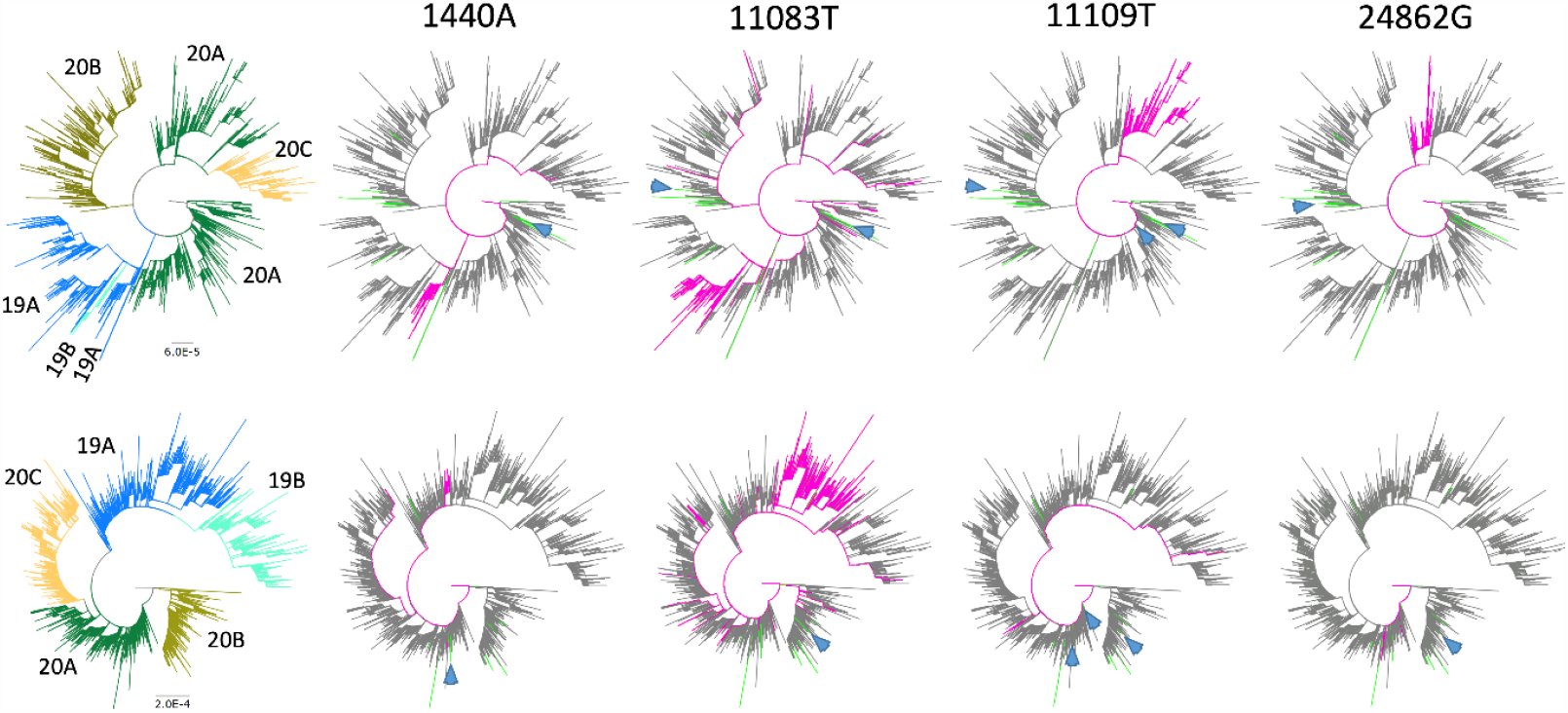
Dutch-Belgian (top) and Global subsample (bottom) phylogenetic trees showing 4 variants present in the sewage samples. Patient sequences containing the mutation are shown in magenta. Lines in green indicate sewage samples sequenced in this study. Clades (19A, 19B, 20A, 20B and 20C) are indicated in colors at the left of the figure. Blue arrows show the consensus sequences (if available) of the sewage samples in which the LFV was detected.

## DISCUSSION

Given the high chemical and biological complexity of wastewater samples, virus concentration and RNA extraction methods are crucial parts of the process to reach enough viral RNA yield for sequencing^29^. In this study, we showed that our method was capable of obtaining complete or near complete genomes from wastewater samples with Ct values of at least 5 or 6 Cts below the limit of detection (LoD) (Ct < 39) and partial genomes for samples with higher Ct values. Therefore, only samples with enough viral RNA can be used to effectively analyze SARS-CoV-2 diversity in sewage samples. In order to increase the percentage of genome covered of the samples, we used a threshold of 10X coverage per position to generate the consensus sequences from Nanopore reads. Based on previous analysis of viral sequencing data, the error rate with this threshold is less than 0.03%^30^. The majority of the mutations (132/145) listed in Supplementary Table S2 have a coverage of at least 30X, which produces an error rate of 1 in 585,000 nucleotides sequenced^30^.

The use of sewage as a tool to understand the epidemiology and diversity of SARS-CoV-2 at a community level offers many advantages over human sampling. Sewage samples are relatively easy to collect, because no invasive sampling is required, there is no sampling bias towards sequences from moderate and severe cases, there are limited ethical issues, and potentially few samples are required to give a picture of the temporal changes of viral infections in the community^28,29^. Nevertheless, comprehensive comparisons with clinical surveillance and other epidemiological approaches are required to determine the extent and limits of using sewage as a surveillance/early warning tool. Furthermore, before the broad use of sewage samples to characterize viral diversity within a population, some obstacles need to be overcome, such as: low viral titers that complicate the retrieval of complete genomes and the distinction of multiple strains within a sample. Here we have used two of the most common NGS technologies (Nanopore and Illumina) to study the diversity of

SARS-CoV-2 found in sewage samples, from the Netherlands and Belgium, and compared these results with the virus diversity found in sequenced clinical samples. In order to evaluate this diversity in a comprehensive fashion, we have used Nextstrain clade classification system because it is based on signature mutations to assign a sequence to a clade (https://nextstrain.org/)^5^, facilitating the association of SNPs or LFV to a particular clade, especially for genome sequences with <75% coverage.

Sewage samples can contain a mixture of SARS-CoV-2 viruses reflecting the multiple viruses circulating within a community. They may also partially reflect the presence of SARS-CoV-2 from animal origin, as SARS-CoV-2 has now been detected in domestic and livestock animals such rabbits, minks, cats and ferrets^31–35^. The analysis of a consensus sequence genome from a wastewater sample may identify the predominant virus strain present in a population, which might be suitable for locations where only 1 or few introductions of closely related viruses have occurred, as it seems to be the case for 2 previous studies in Italy and USA^23,24^. Nonetheless, the consensus genome approach cannot reflect the diversity of the viruses circulating in a population with a high degree of viral diversity. Moreover, in some cases samples containing several diverging strains at significant levels might lead to retrieve artificial consensus genomes that do not represent an existing virus, which seems to be the case for the hCoV-19/env/Netherlands/Amersfoort-92503-N/2020 sequence, where signature mutations of 3 different clades are present at the consensus level. In this study, we could not detect a genome belonging to the least prevalent clades (20C and 19B), despite the circulation of these viruses in the human population in both countries during the same period of time (Figs 2a and b). Although, it is necessary to mention that mutations associated with clades 19B and 20C were found in 2 and 3 samples, respectively (Supplementary Table S1). However, these consensus sequences were either too short or had a mixture of signature mutations that did not allow to confirm whether they belong to clades 20C and 19B by the Nextclade tool. Another reason to explain why we did not find consensus sequences belonging to these clades is the limited number of locations represented on the phylogeny by our sewage sample dataset compared with that of the clinical samples, especially for Belgium (only 2 sequences from sewage).

In depth NGS analysis could help to unravel the diversity of viruses within a complex sample such as wastewater, particularly unbiased sequencing of the sewage virome can give a good picture of the general viral diversity contained in a sample^36^. Nevertheless, the detection of variants of a particular virus in a single sample can be difficult due to the relative low number of reads obtained for each virus type. Targeted amplification of a genome region of the virus taxa of interest is potentially more sensitive and cheaper to perform. Recently, examples of this have been described for enteroviruses, human mastadenoviruses and norovirus^16,37,38^.

In general, for each virus a specific small fragment (< 400 bp) of the genome is amplified and deep-sequenced, then sequencing reads can be aligned and assigned to a particular genotype or serotype, identifying and determining the prevalence of several virus variants within a single wastewater sample^16,37,38^. As the diversity of SARS-CoV-2 is still limited^39^, this approach would not be as useful for this virus because no single small piece of the genome can reliably differentiate between clades or lineages. However, we tried to overcome this issue by using a variant analysis of sewage samples. We showed that some LFVs can be linked to particular clusters or clades within the trees (Fig 3), without the need of a complete genome. Although, in order to confidently determine the presence of a particular clade/cluster within a sample, at least 2 or 3 LFVs associated with such clade/cluster should be present at significant levels. Furthermore, variant analysis can also be used to monitor the prevalence of biological interesting mutations in a population. One of the most interesting is the D614G (or A23403G) mutation in the S glycoprotein, that has been shown to increase infectivity *in vitro* by stabilizing the S1/S2 interaction^40^, and has been associated with higher transmission and mortality rates, although the latter is under debate^41,42^. Unfortunately, the region containing the D614G mutation was not sequenced at high enough coverage to perform a variant analysis in most of the tested samples, and we were not able to find any sample with a mixture of both variants (614D and 614G).

The combination of whole-genome sequencing of clinical samples with epidemiological data has shown to be important for public health decision-making^43^. This type of data can help to identify clusters of infection, occasional new introductions, expansion and decline of circulating strains. The dynamics of virus spread and diversity can vary in each location. It is expected that cities or regions with a high number of visitors have several introductions of the virus, while the opposite is expected in regions with low displacement of people. The use of consensus sequence and LFV analyses in sewage samples can help to swiftly evaluate the diversity of the virus within a particular location and its changes over time. For example, in scenarios where an increase of viral diversity is detected in sewage, suggesting new introductions or the appearance of new clusters of infection, appropriate measures can be taken.

Wastewater can also be used to monitor novel mutations. Our consensus and LFV analyses revealed a total of 57 mutations that were not present in the global database. From these, 51 were found at the consensus level, 8 as LFVs and 2 were common to both analyses. It is possible that these novel mutations were not previously detected due to several reasons: 1) Genetic drift eliminated these variants before they could be further spread and detected; 2) These viruses cause only asymptomatic or mild disease that made them to be less likely to be detected through clinical sampling; 3) They are associated with reduced transmission/replication and did not became fixed in the population; 4) They originate from an unknown animal hosts; 5) They are associated with enhanced enteric shedding/replication; 6) They are associated with intra-host defective genomes. The latter has previously been suggested for the detection of LFVs that generate stop codons in ORF1ab and S genes^44^, and here 4 SNPs that generate stop codons in ORF1ab (Supplementary Table S2); 7) They could also reflect PCR errors during library preparation. Given the relative low concentration of SARS-CoV-2 RNA in some sewage samples, a polymerase mistake during the initial PCR cycles of the library preparation can be amplified and identified as a variant. Phenotypical studies could help to determine the likelihood and biological relevance of some of these novel mutations.

In conclusion, this work illustrates how NGS analysis of wastewater can be used as a tool to approximate the diversity of SARS-CoV-2 viruses circulating in a community. Sequencing of wastewater samples could be a powerful tool to complement clinical surveillance or be used as a standalone procedure in settings where wide clinical sequencing is not feasible.

Additionally, in-depth NGS analysis of wastewater samples can help in assessing changes in virus diversity to determine emergence of epidemiologically or clinically relevant mutations, aiding public health decision making.

## METHODS

### Sample preparation

A total of 55 wastewater (WW) specimens were included in this study. RNA from 7 of these WW samples (all from March 25^th^) were collected, processed and extracted previously by KWR^18^. The other 48 WW specimens were collected as 24 h flow-dependent composite samples and processed as previously described^18^. Briefly, debris of 100 - 200 mL of sewage samples were pelleted and the supernatant concentrated by ultrafiltration in 100 kDa Centricon® Plus-70 centrifugal ultrafilters (Millipore, Amsterdam, The Netherlands). A previously described non-target RNA fragment^19^ was added to sewage concentrates as an internal extraction control. RNA was extracted using the magnetic extraction reagents of the Biomerieux Nuclisens kit (Biomerieux, Amersfoort, The Netherlands) and the semi-automated KingFisher mL (Thermo Scientific, Bleiswijk, The Netherlands) purification system. The RNA was screened for the presence of both SARS-CoV-2 and the internal control by reverse-transcriptase quantitative real-time PCR (RT-qPCR) using the Taqman Fast Virus 1-Step Master Mix (ThermoFisher Scientific, Landsmeer, The Netherlands) with 5 primers/probe sets. Three of these sets target different regions of the SARS-CoV-2 Nucleocapsid (N) gene (N1-N3)^25^, one set targets the envelope (E) gene for all *Sarbecoviruses* ^26^ and the final set targeting the internal control^19^. Only RNA from sewage surveillance samples that had a Ct value below 36 in SARS-CoV-2 specific RT-qPCR assays were further processed for sequencing. RNA extracts from sewage samples were stored at -80 °C.

### Next generation sequencing (NGS) of SARS-CoV-2 genomes by Nanopore and Illumina

SARS-CoV-2 specific multiplex PCR for Nanopore sequencing was performed as described by Oude Munnink, *et al*. (2020)^43^. In short, primers for 89 overlapping amplicons spanning the entire genome were used in 2 PCR pools. The amplicon length was set to 500bp with 75bp overlap between the different amplicons. The used concentrations and primer sequences have been described previously^43^. Libraries were generated using the Oxford Nanopore’s native barcode kits (Catalog numbers: EXP-NBD104, EXP-NBD114 and SQK-LSK109) and sequenced on a R9.4 flow cell multiplexing up to 24 samples per sequence run.

Illumina sequencing was performed as described by Richard, *et al*. (2020)^45^. Amplicons were generated by the SARS-CoV-2 specific multiplex PCR described above for the whole genome Nanopore sequencing. Amplicons were purified with 0.8X AMPure XP beads (Beckman Coulter) and 100 ng of DNA was converted into paired-end Illumina sequencing libraries using the KAPA HyperPlus library preparation kit (Roche), following the manufacturer’s recommendations. Multiplex Adaptors with indexes (KAPA Unique Dual-Indexed Adapters Kit, Roche) were used to enable subsequent sequencing of multiple libraries in a single Illumina V3 MiSeq flowcell (2×300 cycles). Libraries from all re-sequenced samples (both Nanopore and Illumina) were generated from the same RNA as starting material. Consensus sequences with coverage >50% were upload to GISAID (https://www.gisaid.org/), accession IDs: EPI_ISL_539300 - EPI_ISL_539325.

### Nanopore sequence data analysis

The resulting raw sequence data were processed as previously described by Oude Munnink et al. 2020^43^. Briefly, an automated snakemake script^46^ was used to demultiplex fastq raw reads using Porechop (https://github.com/rrwick/Porechop), trim primers using Cutadapt^47^ and perform a reference-based alignment using minimap2 to GISAID sequence EPI_ISL_412973. The run was monitored using RAMPART (https://artic-network.github.io/rampart/). The consensus genome was extracted and positions with a coverage < 10X or <30X were replaced with an “N”. Mutations in the genome were confirmed by manually checking the alignment in Ugene^48^ and homopolymeric regions were manually resolved consulting reference genomes. Based on previous validation studies^30^, mutations with a cut-off of 30X coverage were considered as high quality, whereas mutations with less than 30X coverage were marked as low quality (Supplementary Table S2).

### Illumina Data Filtering, Genome assembly and Variant calling

All the processing, reference-based alignment and variant analysis of the Illumina generated data was performed using a customized workflow on the Galaxy EU server (https://usegalaxy.eu/)^49^. First, raw sequencing reads were filtered using Fastp^50^ to remove adaptor contamination, ambiguous bases (N), low quality reads (Phred score <30) and fragments below the length of 50 nt. For mapping purposes, reads were aligned against the GISAID sequence EPI_ISL_412973 using the default penalty settings of the BWA-MEM^51^.

Reads were re-aligned using the leftalign utility from FreeBayes package^52^. All reads with mapping scores of less than 30 were discarded. Both consensus sequences and variants were generated using iVar^53^. Final consensus sequences (frequency >50%) were constructed using all mapped sequence reads that covered each site at least 5 times and had a minimum quality Phred score of 30. For detection of low-frequency variants (LFV), parameters were set as follows: a minimum coverage of 50X, Phred score >30 and a Minimum frequency threshold of 10%. Manual inspection of the aligned reads was also performed to confirm or dismiss the variant calling in Ugene^48^. Variant positions are given with respect to the Wuhan-Hu-1 strain (Genbank accession number: MN908947)^1^.

### Phylogenetic analysis

Two reference datasets were used to perform the phylogenetic analysis. The first dataset included all Dutch and Belgian full-length SARS-CoV-2 genomes (1544 and 888 sequences, respectively) from GISAID database (https://www.gisaid.org/) publicly available up to the 8^th^ of July 2020. The second dataset is a subsample of all SARS-CoV-2 sequences available in GISAID (https://www.gisaid.org) covering the global diversity of SARS-CoV-2 genomes up to the 1^st^ of June 2020. This global ‘backbone’ dataset contains 2552 subsampled high-quality sequences (full length, with Ns <5%) to include one unique genome per country/state per week. For the maximum-likelihood (ML) trees, only sequences in this study with >75% genome coverage were included in the analysis. Our sequences were aligned with both datasets using MAFFT (https://mafft.cbrc.jp/alignment/server/). The alignment was manually checked for discrepancies and the ends were trimmed, after which IQ-TREE^54^ was used to perform a ML phylogenetic analysis under the GTR + F + R3 model for the Global subsample and the GTR + F + R2 model for the Dutch-Belgian dataset as the best predicted models using the ultrafast bootstrap option with 1,000 replicates. The phylogenetic trees were visualized using Figtree v1.4.4 (http://tree.bio.ed.ac.uk/software/figtree/). Clades were assigned by using the Nextclade tool within Nextstrain (https://clades.nextstrain.org/).

## Data Availability

Consensus sequences with coverage >50% were upload to GISAID (https://www.gisaid.org/), accession IDs: EPI_ISL_539300 - EPI_ISL_539325.

## ACKNOWLEDGEMENTS

This work was supported by the European Union’s Horizon 2020 grants VEO (grant nr 874735) and METASTAVA (grant nr 773830), the Erasmus MC foundation and the Adessium Foundation. The authors thank the Water Authorities in the Netherlands (Aa en Maas, Amstel Gooi en Vecht, Delfland, De Dommel, Fryslan, Hollands Noorderkwartier, Stichtse Rijnlanden, Vallei en Veluwe, Evides, Waternet), and Belgium (Aquafin, De Watergroep) for the provision of the sewage samples. We gratefully acknowledge the authors, originating and submitting laboratories of the global sequences from GISAID’s EpiCoV™ Database^3^ on which this research is based.

## AUTHOR CONTRIBUTIONS

RIL, MPGK and MdG wrote the report. GE, LH and GM set up sample collection. GE, LH, CS generated sequence data. RIL, DN, MK, BOM, LL, SL and MdG were involved in data analysis and interpretation. RIL, FA, GM, MPGK and MdG designed the study. All authors provided critical feedback and contributed to manuscript editing.

## COMPETING INTERESTS

The authors declare no competing interests

## SUPPLEMENTARY MATERIAL

**Supplementary Table S1**. Summary of all SNPs detected in wastewater samples at the consensus level.

**Supplementary Table S2**. Summary of the mutations detected in the genome consensus sequences from wastewater samples determined by both Nanopore and Illumina sequencing. Coverage is indicated as Illumina if the mutation was found only in Illumina sequencing, High if the Nanopore coverage was >30X and Low if the coverage was between 10X and 30X.

**Supplementary Fig. S1**. RT-qPCR cycle threshold (Ct) of SARS-CoV-2 RNA in sewage as determined by the N gene (N1-N3) and the E gene PCR assays against the percentage of the genome covered by Illumina reads.

**Supplementary Fig. S2**. Fully extended phylogenetic tree of SARS-CoV-2 genome consensus sequences detected in sewage samples in the Netherlands and Belgium within the Dutch-Belgian dataset.

**Supplementary Fig. S3**. Fully extended phylogenetic tree of SARS-CoV-2 genome consensus sequences detected in sewage samples in the Netherlands and Belgium within the global subsample dataset.

## Notes

### Competing Interest Statement

The authors have declared no competing interest.

### Author Declarations

Not applicable. Patient-derived viral sequences were obtained from GISAID database (https://www.gisaid.org/). Our samples were obtained from community wastewaters, which do not require an approval for their use.

